# Preliminary exploration of facial electromyographic indices of pain in people undergoing hand surgery

**DOI:** 10.1101/2022.11.23.22282655

**Authors:** Filip Panchevski, Ifigeneia Mavridou, Hristijan Gjoreski, Martin Gjoreski, Ivana Kiprijanovska, Simon Stankoski, Charles Nduka, John Broulidakis

## Abstract

Assessing pain levels in real-world conditions, such as during active surgery, can be challenging. Self-reports, often considered globally as ‘ground-truth’ can be unreliable, episodic and ill-suited to routine monitoring or use with non-verbal patients. Lately, physiological measurements have been explored as an objective method for assessing the symptoms of pain increase on the body. We investigated the effects of pain (high pain) using facial mask – emteqPRO, equipped with seven facial electromyographic (fEMG) sensors.

Our aims were to: (i) investigate the efficacy of continuous physiological monitoring within surgery at a hospital environment, and (ii) to produce initial findings and show how pain increase affects the data from fEMG sensors.

## Methods

### 1. Subjects

Forty adult volunteers’ outpatients from Queen Victoria Hospital were selected and screened to take part in this study. Eligibility criteria excluded history of facial neuromuscular disease - Bell’s palsy, chronic pain, or previous facial surgery were selected. All participants have had received superficial trauma to the hand only. From those, 9 random datasets were selected and used for this preliminary analysis.

### 2. Experimental

The procedure of the data collection was two-fold. Upon receiving confirmation that the participant was suitable to be part of the data collection, the first step was taking a five-minute baseline reading of the sensors while the participant was incited to remain relaxed. Afterwards, the second part of the recoding took place when the medical procedure started. In order to not interfere with the operation, the surgeon was instructed to carry out the procedure as if there was no recording, which lasted at most 30 minutes. The recordings consist of sensor data from the facial muscles around the eyes and the corrugator, collected using a wearable biometric device – emteqPRO. The evaluation of the subjective pain felt was reported by the participants on a scale from 0-10 every 30 seconds for the duration of the operation.

### 3. Self-reports and data annotation

Participants reported their subjective pain levels on a scale from 0 (no pain) to 10 (worst pain), similar as in the visual analog scale (VAS)[3]. During the operation, a video with audio reminded the participant to report their pain scores every 30.8 seconds. Those scores were transcribed and used in the analysis.

### 4. Electromyographic measurement

The emteqPRO is a medical-grade wearable mask with integrated physiological sensors that detects the symptoms of emotional changes. It comprises photoplethysmographic (PPG) sensor, 7 facial electromyographic (fEMG) sensors, and an Inertial Measurement Unit (IMU) sensor. This way, it can detect muscle activations from 7 locations: the zygomatic, corrugator, frontalis, and orbicularis muscles. Different size side-inserts were used for participants based on their face size and shape, to ensure maximum sensor-to-skin contact during the data collection. The provided SuperVision monitoring software [4] was used during initial sensor set-up.

### 5. Data processing and analysis

The data streams from the EMG sensors were collected as multiple time series. The frequency of the processed amplitude of the fEMG sensors was set to 50Hz. Two separate data segments were extracted in order to compare the two conditions (Baseline and Pain; low and high level of pain). The first being a one-minute segment from the baseline with the lowest VAS score, and the second segment taken from the data collected whilst the participant was undergoing the medical procedure with the highest VAS score. As a starting point we analysed and calculated average amplitude and conductance for each one of the EMG sensors.

## Results

Table 1 presents the results. In comparison to a pre-operative baseline, the operative elicited increased activity in all facial muscles, but a pronounced difference can be observed from the fEMG sensors covering the zygomatic muscle region. Indeed, this difference was found significant between the base and the pain condition for both the right and left zygomaticus sensors when tested using Wilcoxon paired t-tests (Left zygomaticus: Z=-2.192, p=0.028, Right Zygomaticus: Z=-2.521, p=0.012).

**Table 1.** Average amplitude for each of the EMG sensors.

|  | Base | Pain | Difference (Pain–Base) |
| --- | --- | --- | --- |
| VAS score | 1.0 | 4.778 | 3.778 |
| Right Frontalis | 0.0000047 | 0.0000037 | -0.0000010 |
| Right Zygomaticus | 0.0000018 | 0.0000054 | 0.0000035 |
| Right Orbicularis | 0.0000031 | 0.0000055 | 0.0000025 |
| Center Corrugator | 0.0000023 | 0.0000029 | 0.0000006 |
| Left Orbicularis | 0.0000032 | 0.0000064 | 0.0000032 |
| Left Zygomaticus | 0.0000021 | 0.0000052 | 0.0000031 |
| Left Frontalis | 0.0000031 | 0.0000032 | 0.0000000 |

## Conclusions

The analysis of the data from the participants undergoing hand surgery, showed that there is a significant difference between the fEMG muscle activations when the participant is not experiencing pain compared to a high pain condition. The difference was significant for the zygomaticus (left and right) and highly pronounced but not significant for the corrugator sensors. This initial analysis is a good basis to further explore data predictors of pain and potentially develop an automatic pain estimation algorithm using the data from fEMG sensors. In future work, we plan to include additional data features including heart-rate dynamics.

## Data Availability

All data produced in the present study are available upon reasonable request to the authors.

https://www.emteqlabs.com/

## Ethical Permissions

The study was reviewed and approved by the South Central - Oxford C Research Ethics Committee (19/SC/0274).

## Relevance for Patient Care

Since self-reports, can be unreliable, episodic and ill-suited for use with non-verbal patients, providing a solution that can objectively measure pain changes using physiological signal instead could prove to be a very useful assistive technology for continuous monitoring in clinical environments.

## References

[1] Governo, R., Eden-Green, B., Dawes, T., Mavridou, I., Giles, J., Rosten, C., … & Nduka, C. (2020). Evaluation of facial electromyographic pain responses in healthy participants. Pain Management, 10(6), 399–410.

[2] Gjoreski, H., I. Mavridou, I., Fatoorechi, M., Kiprijanovska, I., Gjoreski, M., Cox, G. and Nduka, C., 2021, September. emteqPRO: Face-mounted Mask for Emotion Recognition and Affective Computing. In Adjunct Proceedings of the 2021 ACM International Joint Conference on Pervasive and Ubiquitous Computing and Proceedings of the 2021 ACM International Symposium on Wearable Computers (pp. 23–25).

[3] Jamison, R.N., Gracely, R.H., Raymond, S.A., Levine, J.G., Marino, B., Herrmann, T.J., Daly, M., Fram, D. and Katz, N.P., 2002. Comparative study of electronic vs. paper VAS ratings: a randomized, crossover trial using healthy volunteers. Pain, 99(1-2), pp.341–347.

[4] Gnacek, M., Broulidakis, J., Mavridou, I., Fatoorechi, M., Seiss, E., Kostoulas, T., Balaguer-Ballester, E., Rosten, C. and Nduka, C., 2022. EmteqPRO-Fully Integrated Biometric Sensing Array for Non-Invasive Biomedical Research in Virtual Reality. Frontiers in Virtual Reality, p.3.

